# A 21L/BA.2-21K/BA.1 “MixOmicron” SARS-CoV-2 hybrid undetected by qPCR that screen for variant in routine diagnosis

**DOI:** 10.1101/2022.03.28.22273010

**Authors:** Philippe Colson, Jeremy Delerce, Elise Marion-Paris, Jean-Christophe Lagier, Anthony Levasseur, Pierre-Edouard Fournier, Bernard La Scola, Didier Raoult

## Abstract

Among the multiple SARS-CoV-2 variants identified since summer 2020, several have co-circulated, creating opportunities for coinfections and potentially genetic recombinations that are common in coronaviruses. Viral recombinants are indeed beginning to be reported more frequently. Here, we describe a new SARS-CoV-2 recombinant genome that is mostly that of a Omicron 21L/BA.2 variant but with a 3’ tip originating from a Omicron 21K/BA.1 variant. Two such genomes were obtained in our institute from adults sampled in February 2022 in university hospitals of Marseille, southern France, by next-generation sequencing carried out with the Illumina or Nanopore technologies. The recombination site was located between nucleotides 26,858-27,382. In the two genomic assemblies, mean sequencing depth at mutation-harboring positions was 271 and 1,362 reads and mean prevalence of the majoritary nucleotide was 99.3±2.2% and 98.8±1.6%, respectively. Phylogeny generated trees with slightly different topologies according to whether genomes were depleted or not of the 3’ tip. This 3’ terminal end brought in the Omicron 21L/BA.2 genome a short transposable element of 41 nucleotides named S2m that is present in most SARS-CoV-2 except a few variants among which the Omicron 21L/BA.2 variant and may be involved in virulence. Importantly, this recombinant is not detected by currently used qPCR that screen for variants in routine diagnosis. The present observation emphasizes the need to survey closely the genetic pathways of SARS-CoV-2 variability by whole genome sequencing, and it could contribute to gain a better understanding of factors that lead to observed differences between epidemic potentials of the different variants.

## INTRODUCTION

Multiple SARS-CoV-2 variants have been identified since summer 2020 (Lemey et al., 2021; Colson et al., 2022a; Hodcroft et al., 2021; Harvey et al., 2021). There were periods during which two distinct variants co-circulated with a crossing of their incidence when an old variant was vanishing and a new one was rising. Substantial rates of co-incidence over periods of several weeks have thus been reported recently worldwide for the Delta [WHO denomination (https://www.who.int/fr/activities/tracking-SARS-CoV-2-variants)]/21J [Nextclade classification (Aksamentov et al., 2021) (https://nextstrain.org/)] and Omicron 21K/BA.1 [Pangolin classification (Rambaut et al., 2020) (https://cov-lineages.org/resources/pangolin.html)] variants, and thereafter, to a lesser extent, for the Omicron BA.1 and BA.2 variants (https://covariants.org/per-country) (Hodcroft, 2021; Hadfield et al., 2018), including in our geographical area (Figure 1). This created the opportunity for coinfections (Rockett et al., 2022; Hosch et al., 2022; Bolze et al., 2022; Belen Pisano et al., 2022), and consequently for homologous genetic recombinations, which constitute a major and very common mechanism of evolution in viruses (Bentley and Evans, 2018). Such genetic recombinations are extremely frequent for viruses of family *Coronaviridae*, and they have already been identified for endemic human coronaviruses (Lai, 1996; Zhang et al., 2015; So et al., 2019; Gribble et al., 2020). Regarding SARS-CoV-2, the occurrence of recombinations has been reported or suspected (Yi, 2019; Yeh and Contreras, 2020; Haddad et al., 2021; Ignatieva et al., 2021; Jackson et al., 2021; Taghizadeh et al., 2021; Varabyou et al., 2021; Kreier, 2022; Wertheim et al., 2022; He et al., 2022; Sekizuka et al., 2022; Colson et al., 2022b; Lacek et al., 2022; Lohrasbi-Nejad, 2022; Bolze et al., 2022; Ou et al., 2022; Belen Pisano et al., 2022; Burel et al., 2022). Very recently, we described the identification and culture of two SARS-CoV-2 recombinants, one between the B.1.160 and Alpha/20I variants in a patient chronically-infected with SARS-CoV-2 (Burel et al., 2022), and another between the Delta/21J AY.4 and Omicron 21K/BA.1 variants in patients infected approximately 10 weeks after the start of the period of co-detection of these two variants in our geographical area (Colson et al., 2022b). Here, we describe a new hybrid genome, which consists of a Omicron 21L/BA.2 genome whose 3’ tip originates from a Omicron 21K/BA.1.

**Figure 1.**
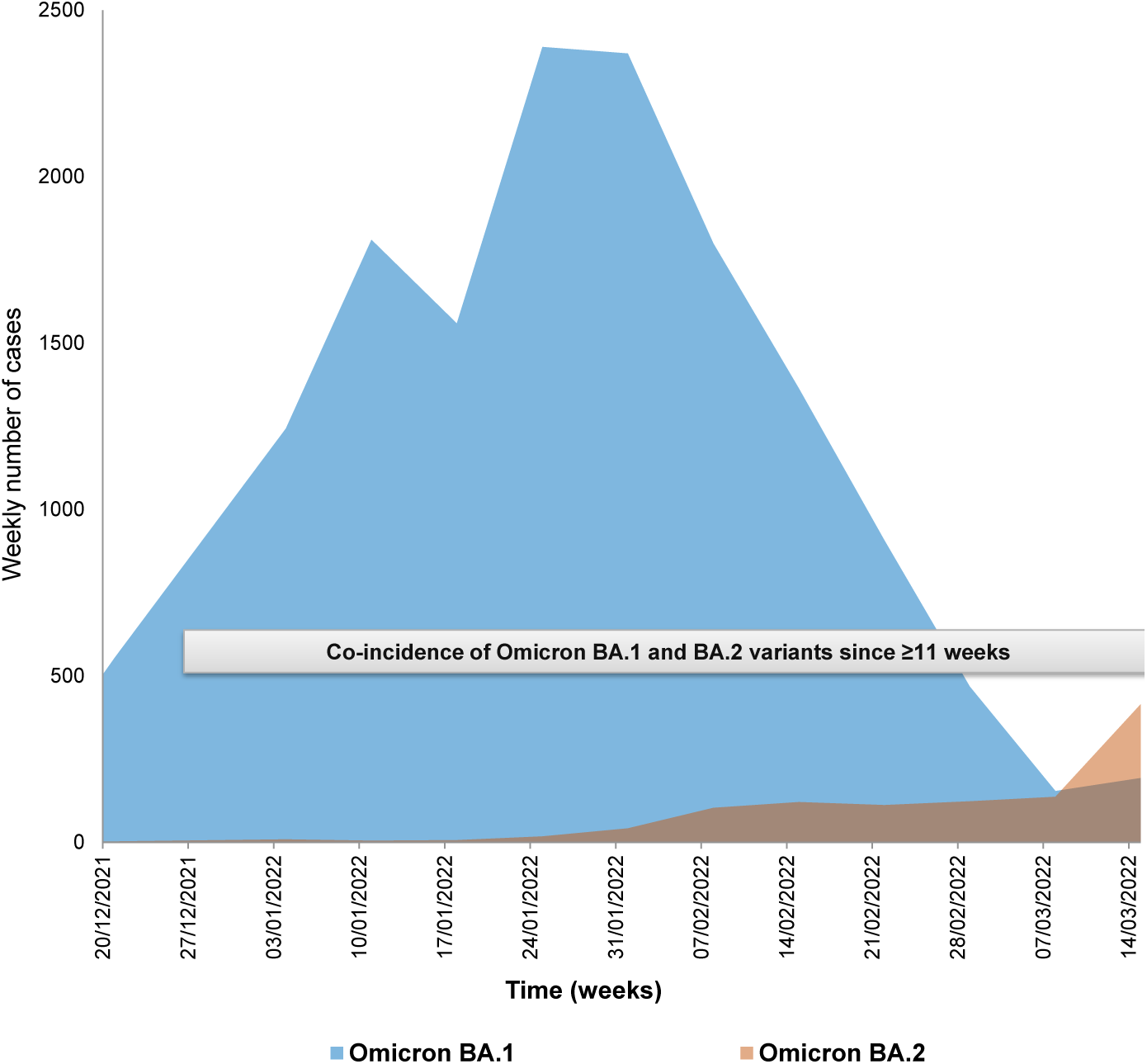
Incidence and co-incidence of Omicron 21K/BA.1 and Omicron 21L/BA.2 variants among cases diagnosed in our institute.

## MATERIALS AND METHODS

Nasopharyngeal samples were tested for the presence of SARS-CoV-2 RNA by real-time reverse transcription-PCR (qPCR) using the BGI real-time fluorescent RT-PCR assay (BGI Genomics, Shanghai Fosun Long March Medical Science Co., Ltd., Shenzhen, China), as previously described (Burel et al., 2022). Subsequently, qPCR assays that screen for SARS-CoV-2 variants were carried out with the detection of mutations among which spike substitution K417N (Thermo Fisher Scientific, Waltham, USA) and the targeting of viral genes S (spike), N (nucleocapsid) and ORF1 with the TaqPath COVID-19 kit (Thermo Fisher Scientific), as previously reported (Colson et al., 2022a; Colson et al., 2022b).

SARS-CoV-2 genomes were obtained and analyzed as described previously (Colson et al., 2022a; Colson et al., 2022b). Briefly, next-generation sequencing was carried out with the Illumina COVID-seq protocol on the NovaSeq 6000 instrument (Illumina Inc., San Diego, CA, USA), or with the Oxford Nanopore technology (ONT) on a GridION instrument (Oxford Nanopore Technologies Ltd., Oxford, UK) following multiplex PCR amplification with the ARTIC nCoV-2019 Amplicon Panel v4.1 of primers (IDT, Coralville, IA, USA) and the ARTIC procedure (https://artic.network/).

Sequence read processing and genome analysis were performed as described previously (Colson et al., 2022a; Colson et al., 2022b). Briefly, for Illumina NovaSeq reads, base calling was carried out using the Dragen Bcl Convert pipeline [v3.9.3; https://emea.support.illumina.com/sequencing/sequencing_software/bcl-convert.html (Illumina Inc.)], mapping was carried out using the bwa-mem2 tool (v2.2.1; https://github.com/bwa-mem2/bwa-mem2) on the Wuhan-Hu-1 isolate genome (GenBank accession no. NC_045512.2) before cleaning with the SAMtools program (v. 1.13; https://www.htslib.org/) (Danecek et al., 2021). Variant calling was performed using FreeBayes (v1.3.5; https://github.com/freebayes/freebayes) (Garrison et al., 2012), and consensus genomes were built with the Bcftools program (v1.13; https://samtools.github.io/bcftools/bcftools.html). ONT reads were processed with the ARTIC-nCoV-bioinformaticsSOP pipeline (v1.1.0; https://github.com/artic-network/fieldbioinformatics). Nucleotide and amino acid changes compared to the Wuhan-Hu-1 isolate genome were determined using the Nextclade tool (https://clades.nextstrain.org/) (Hadfield et al., 2018; Aksamentov et al., 2021). Nextstrain clades and Pangolin lineages were identified with the Nextclade web application (https://clades.nextstrain.org/) (Hadfield et al., 2018; Aksamentov et al., 2021) and the Pangolin tool (https://cov-lineages.org/pangolin.html) (Rambaut et al., 2020), respectively.

Phylogenetic analyses were performed the MEGA X software (v10.2.5; https://www.megasoftware.net/) (Kumar et al., 2018) following sequence alignment with MAFFT (https://mafft.cbrc.jp/alignment/server/) (Katoh et al., 2002), and trees were visualized with MEGA X. We built two separate trees, a first one for the whole recombinant genome and a second one for its part originating from an Omicron 21L/BA.2 variant. The 10 genomes the most similar to these sequences among genomes of the Omicron 21L/BA.2 and 21K/BA.1 variants of the sequence database of our institute were selected by a BLAST search (Altschul et al., 1990) then incorporated in the phylogenies together with the sequence of the Wuhan-Hu-1 isolate.

Genome sequences obtained and analyzed here were deposited in the NCBI GenBank nucleotide sequence database (https://www.ncbi.nlm.nih.gov/genbank/) (Sayers et al., 2022) (Accession no. OM993515 and OM993473), on the IHU Méditerranée Infection website (https://www.mediterranee-infection.com/sars-cov-2-recombinant/) (IHUCOVID-063942 and IHUCOVID-068136), and in the GISAID database (https://www.gisaid.org/) (Elbe et al., 2017; Alm et al., 2020) (EPI_ISL_10843457, EPI_ISL_10047082).

## RESULTS

The recombinant genomes were obtained from two adult patients sampled in February 2022 in the university hospitals of Marseille, southern France. Cycle threshold values (Ct) of the diagnosis qPCR performed in our institute were 13 and 19. The TaqPath COVID-19 assay showed positivity for all targeted genes including the S gene, and the spike K417N mutation was detected, which led to suspect an infection with the Omicron 21L/BA.2 variant. However, next-generation sequencing performed in our laboratory allowed obtaining SARS-CoV-2 Omicron 21L/BA.2-21K/BA.1 genomic hybrid forms. These hybrid genomes have a Omicron 21L/BA.2 backbone but their approximately 2,500-3,000 nucleotide-long 3’ terminal region is that of a Omicron 21K/BA.1 (Figure 2). The recombination site is located between nucleotides 26,858 and 27,382 (in reference to the genome of the Wuhan-Hu-1 isolate). This 3’ terminal end in the genomes of the Omicron BA.1 variant does not harbor any signature mutations, and therefore a “gain” of mutation could not be observed here. However, we noted the presence and integrity of a sequence corresponding to a short transposable element of 41 nucleotides named S2m (Tengs et al., 2021), which is present in the Omicron 21K/BA.1 variant but is truncated of 26 nucleotides in its central part in the Omicron 21L/BA.2 variant. Current strategies of variant screening by qPCR therefore fail to detect this recombinant as they target mostly mutations in the spike gene, or other genomic regions that do not allow identifying it either. In the two genomic assemblies, the mean (±standard deviation) sequencing depth at positions harboring mutations relatively to the genome of the Wuhan-Hu-1 isolate was 271±164 and 1,362±1,146 reads and the mean prevalence of the majoritary nucleotide was 99.3±2.2% and 98.8±1.6%, respectively, which rules out the concommitant presence of the sequences of two variants in the samples, either due to co-infection or to contamination.

**Figure 2.**
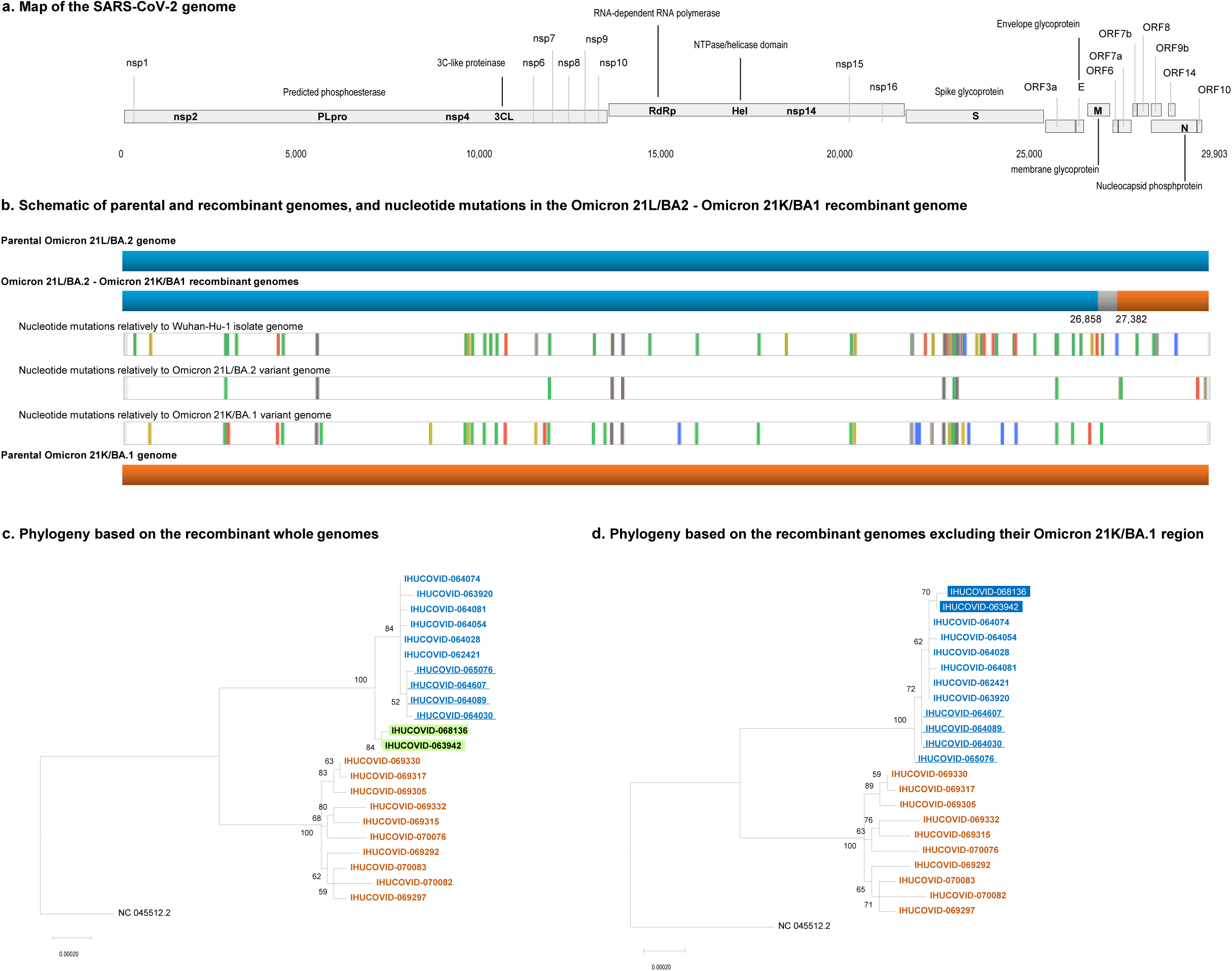
Schematic of the SARS-CoV-2 Omicron 21L/BA.2-21K/BA.1 recombinant genomes (a, b) and phylogeny reconstruction based on SARS-CoV-2 Omicron 21L/BA.2 and Omicron 21K/BA.1 genomes (c, d). a. Map of the SARS-CoV-2 genome. b. Schematic representation of parental and recombinant genomes and mutations in the recombinant genomes. Adapted from screenshots of the nextclade web application output (https://clades.nextstrain.org) (Aksamentov et al., 2021; Hadfield et al., 2018). Color codes for nucleotide mutations are as follows: Green: U; yellow: G; blue: C; red: A; light grey: deletions; dark grey: regions uncovered by sequencing reads. c, d. Phylogeny reconstructions based on the whole genomes (c) or only on the region of these genomes that corresponds to the region of the recombinant genome that originates from the Omicron 21L/BA.2 variant.

The two recombinant genomes harbor 65 mutations relatively to the genome of the Wuhan-Hu-1 isolate, including 3 or 4 that are not signature mutations of the parental Omicron 21L/BA.2 or Omicron 21K/BA.1 genomes (Table 1). The spike genes of the two genomes only harbor signature mutations of the Omicron 21L/BA.2 variant, apart from a synonymous mutation in one genome. Besides, these two genomes differ between each other by a single mutation in the ORF1 gene (C2790U; T841I).

**Table 1.**
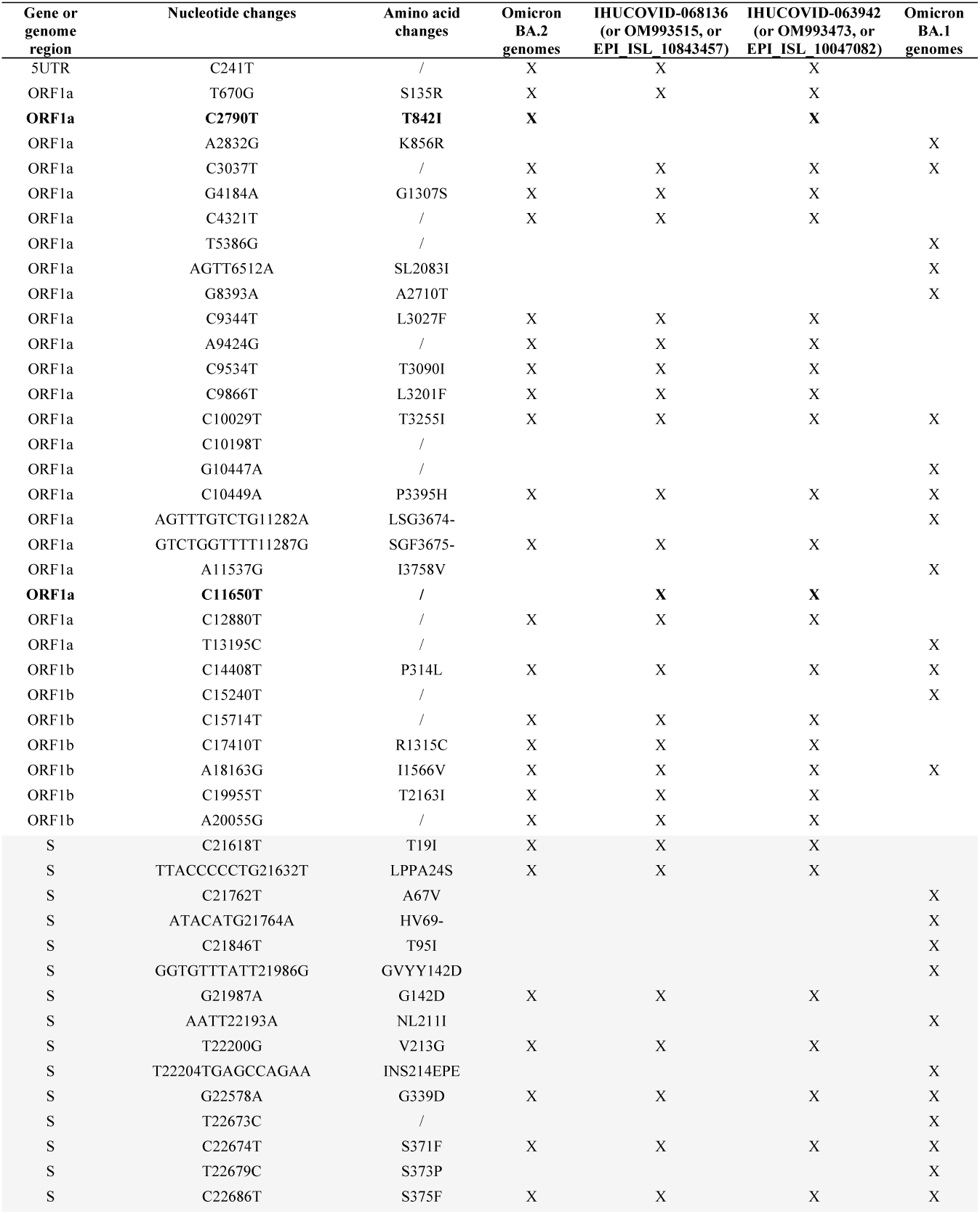
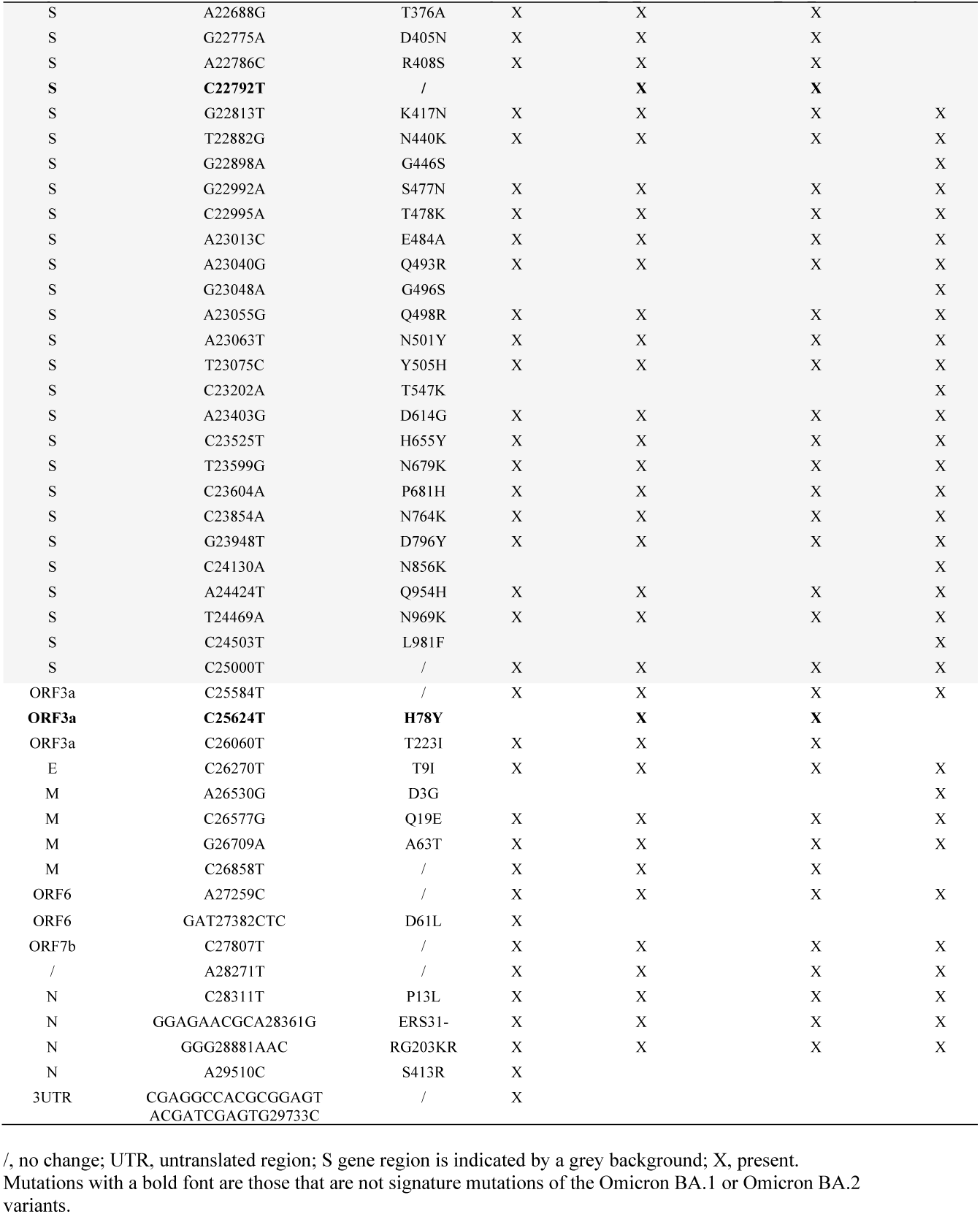
Nucleotide and amino acid changes in the MixOmicron recombinant according to their presence/absence in the Omicron 21L/BA.2 and Omicron 21K/BA.1 variants.

The two phylogenetic trees that were built, based either on the whole genomes or only on the region of these genomes that originate from the Omicron 21L/BA.2 variant in the recombinants, did not show exactly the same topology (Figure 2c, d). The trees incorporated the 10 genomes of Omicron 21L/BA.2 and 21K/BA.1 variants from the sequence database of our institute that were the most similar to the sequences of the recombinants. In both trees, sequences from the recombinants were clustered with sequences of the Omicron 21L/BA.2 variant. Nonetheless, whole recombinant genomes were clustered separately, apart from their best hits, in the Omicron 21L/BA.2 clade that encompassed two sister groups. In contrast, the two partial recombinant genomes were clustered together but nested inside the Omicron 21L/BA.2 clade.

## DISCUSSION

In the present work we show a recombination between Omicron BA.2 and BA.1 variant viruses. In two previous studies, 16 (0.006%) of 279,000 genomes and 1,175 (0.2%) of 537,360 genomes were identified as being recombinants (Jackson et al., 2021; VanInsberghe et al., 2021). In addition, it was deemed that up to 5% of SARS-CoV-2 that circulated in the USA and UK might have been recombinants (VanInsberghe et al., 2021), and estimated that ≈2.7% of sequenced SARS-CoV-2 genomes might have recombinant ancestry (Turkahia et al., 2021). As a matter of fact, cases of detection of recombinant genomes are increasingly reported in 2022 (Wertheim et al., 2022; Sekizuka et al., 2022; Colson et al., 2022b; Lacek et al., 2022; Bolze et al., 2022; Ou et al., 2022; Belen Pisano et al., 2022; Burel et al., 2022). Such recombinants are all the more likely to be generated when different variants co-circulate with high incidence levels. In our geographical area, >15,000 infections with the Omicron 21K/BA.1 variant and >1,000 infections with the Omicron 21L/BA.2 variant were diagnosed over the same period of 11 weeks between late December and mid-March (Figure 1). Moreover, recombinants becomes more easily identifiable during bioinformatic analyses due to the accumulation over time of mutations along SARS-CoV-2 genomes, at intervals of increasingly reduced size. The two phylogeny reconstructions based on whole genomes or on their region that originate from the Omicron 21L/BA.2 variant in the recombinants exhibited slightly different topologies. This emphasizes that phylogenetic analyses do not accurately handle genomes that are hybrids of sequences with different evolutionary histories, which prompts building separate trees for sequences of different origins in the case of recombinants.

The spike gene of the recombinant virus identified here is typical of that of the Omicron 21L/BA.2 variant, which suggests similar phenotypic features regarding immune escape (Yu et al., 2022; Iketani et al., 2022). However, the acquisition by a Omicron 21L/BA.2 genome of the 3’ terminal part of a Omicron 21K/BA.1 genome is of very particular interest as this Omicron 21K/BA.1 fragment contains a short transposable element named S2m. S2m is a 41-nucleotide long stem loop motif that is present in four different families of positive-sense single-stranded RNA viruses (Robertson et al., 1998; Imperatore et al., 2022) and also shows high levels of similarity with sequences of insects (Tengs et al., 2021). This element is present in sarbecoviruses and in most of the SARS-CoV-2 genomes. It was proposed to be involved in RNA interference pathways, in hijacking of host protein synthesis, and in RNA recombination events (Imperatore et al., 2022; Gallaher et al., 2022), and it could have initiated viral infection and pathogenicity in various animal hosts. For instance, the s2m element was reported to interact with cellular miRNA-1307-3p in humans, being putatively able to manipulate the host immune response. Moreover, in SARS-CoV-2, S2m is absent or truncated in a few variants including the Eta (B.1.525), Iota (B.1.526) and B.1.640.1 lineages, which all had a low epidemic spread; and it is also truncated in the Omicron 21L/BA.2 variant. Taken together these data suggest that S2m could be considered as the equivalent of a virulence factor. The consequence of the S2m acquisition by an Omicron 21L/BA.2 genome as reported here is unknown. A possibility would be a gain in transmissibility leading to a larger epidemic, which should be investigated by genotypic surveillance among SARS-CoV-2 diagnoses and phenotypic *in vitro* experiments. Thus, the present observation may contribute to gain a better understanding of factors that enhance SARS-CoV-2 spread and lead to the observed differences between the epidemic potential of the variants (Tao et al., 2021; Campbell et al., 2021).

Finally, the increasing identification of recombinant SARS-CoV-2 genomes worldwide highlights the unpredictable nature of the genetic variability of this virus. The recombinant described here is not detected by current strategies that screen for variants in routine diagnosis by qPCR. This emphasizes the interest of the most exhaustive whole-genome based surveillance possible to allow deciphering the genetic pathways of the variability and investigating their phenotypic consequences regarding transmissibility, clinical severity, and escape from neutralizing antibodies.

## Data Availability

Genome sequences generated and analyzed in the present study are available from the NCBI GenBank nucleotide sequence database (https://www.ncbi.nlm.nih.gov/genbank/) (Accession no. OM993515 and OM993473), from the IHU Mediterranee Infection website (https://www.mediterranee-infection.com/sars-cov-2-recombinant/) (IHUCOVID-063942 and IHUCOVID-068136), and from the GISAID database (https://www.gisaid.org/) (EPI_ISL_10843457, EPI_ISL_10047082).

https://www.mediterranee-infection.com/sars-cov-2-recombinant/

https://www.gisaid.org/

## Acknowledgments

We are very grateful to Raphael Tola, Ludivine Bréchard, and Claudia Andrieu for their technical help.

## Author contributions

Study conception and design: Philippe Colson, Pierre-Edouard Fournier, Bernard La Scola, Didier Raoult. Materials, data and analysis tools: Philippe Colson, Jeremy Delerce, Elise Marion-Paris, Jean-Christophe Lagier, Anthony Levasseur. Data analyses: Philippe Colson, Jeremy Delerce, Elise Marion-Paris, Jean-Christophe Lagier, Anthony Levasseur, Pierre-Edouard Fournier, Bernard La Scola, Didier Raoult. Writing of the first draft of the manuscript: Philippe Colson, Pierre-Edouard Fournier, Bernard La Scola, Didier Raoult. All authors read, commented on, and approved the final manuscript.

## Data availability

Genome sequences generated and analyzed in the present study are available from the NCBI GenBank nucleotide sequence database (https://www.ncbi.nlm.nih.gov/genbank/) (Sayers et al., 2022) (Accession no. OM993515 and OM993473), from the IHU Méditerranée Infection website (https://www.mediterranee-infection.com/sars-cov-2-recombinant/) (IHUCOVID-063942 and IHUCOVID-068136), and from the GISAID database (https://www.gisaid.org/) (Elbe et al., 2017; Alm et al., 2020) (EPI_ISL_10843457, EPI_ISL_10047082).

## Ethics

This study has been approved by the ethics committee of the University Hospital Institute Méditerranée Infection (No. 2022-008). Access to the patients’ biological and registry data issued from the hospital information system was approved by the data protection committee of Assistance Publique-Hôpitaux de Marseille (APHM) and was recorded in the European General Data Protection Regulation registry under number RGPD/APHM 2019-73.

